# Incidence and Predictor of Ocular Hypertension after Intravitreal Injection of Bevacizumab among Patient Attended at KCMC Hospital 2023-2024

**DOI:** 10.1101/2025.03.21.25324412

**Authors:** John Andambike. Ambakisye, Furahin Godfrey Mndeme, William Makupa

## Abstract

**Objective:** This study aimed to evaluate the incidence, patterns, and determinants of ocular hypertension (OH) following Bevacizumab Intravitreal injections for various retinal diseases at KCMC Hospital, Tanzania.

**Methods:** A prospective cohort study was conducted from August 2023 to July 2024, involving 120 participants. OH was defined as an intraocular pressure (IOP) >21 mmHg or an increase >5 mmHg from baseline. Data on demographics, injection history, ocular conditions, and systemic factors were collected. IOP was measured at baseline, immediately post-injection, and at six-week intervals during follow-up. Paired t-tests compared mean IOP differences, Nelson Allan estimator curves assessed cumulative OH risk, and Poisson regression identified associated factors.

**Results:** Participants’ median age was 62 years, with diabetic macular edema (52.5%) being the most common indication. OH incidence was 15%, significantly associated with the number of injections (adjusted hazard ratio [AHR] 2.17, 95% CI 1.56-3.16, p < 0.001) and history of YAG laser capsulotomy (AHR 0.33, 95% CI 0.12-0.88, p = 0.028). Temporary post-injection IOP spikes normalized within 60 minutes.

**Conclusion:** The study revealed a higher incidence of ocular hypertension following Bevacizumab injections compared to other studies. Significant factors included injection frequency and a history of YAG laser capsulotomy, with repeated injections leading to delayed normalization of intraocular pressure and increased spikes during subsequent visits.

## INTRODUCTION

In recent years anti-VEGF agents such as Bevacizumab (Avastin) have revolutionized the management of various retinal diseases worldwide, including diabetic retinopathy, age-related macular degeneration (AMD), and retinal vein occlusions. Bevacizumab, a recombinant humanized monoclonal IgG1 inhibits VEGF-A which lead to suppression of pathological angiogenesis and vascular leakage, thereby preserving visual function(Kabunga *et al.*, 2022).

While the therapeutic benefits of Bevacizumab are well-documented, Intravitreal anti-VEGF injections are associated with potential complications, notably ocular hypertension (OHT), which can lead to glaucomatous damage if left unmanaged(3). OHT refers to a condition where the intraocular pressure (IOP) is elevated above the normal range (typically >21 mmHg) without any detectable optic nerve damage and visual field loss.

The exact mechanisms by which Bevacizumab causes OHT are not entirely understood, but several theories exist. VEGF inhibition might affect the regulation of aqueous humor dynamics, potentially leading to increased production or decreased outflow. Intravitreal injections can induce an inflammatory response, causing trabecular meshwork dysfunction, leading to reduced outflow and increased IOP. The injected fluid itself could cause a transient increase in IOP. Additionally, VEGF plays a role in maintaining normal ocular blood flow; its inhibition might disrupt this balance, or the TM being mechanically blocked by protein clumps or foreign particles(4).

The global burden of ocular hypertension following Intravitreal Bevacizumab injections varies significantly across regions. Studies have shown that the incidence ranges from 4% to 15%, depending on patient demographics, underlying conditions, and the number of injections received(4)

Previous studies have explored risk factors influencing OHT, such as the number of injections pre-existing glaucoma and history of YAG capsulotomy with mixed and inconclusive results(5–8).

Transient intraocular pressure (IOP) spikes immediately following Bevacizumab Intravitreal injections have been reported in several studies; however, the incidence of these spikes varies significantly across different research investigations(9–11).

In Africa, comprehensive data is scarce; however, available studies suggest that the prevalence might be comparable to global trends. Sub-Saharan Africa and East Africa face additional challenges, such as limited access to advanced ophthalmic care and follow-up, which could affect the management and outcomes of patients receiving Intravitreal injections. In Tanzania, specific data on the prevalence, pattern and factor associated with ocular hypertension post-Bevacizumab injection is limited, highlighting a significant gap in regional research. This study aim to evaluate the incidence and predictor of ocular hypertension at KCMC hospital in northern part of Tanzania, this will help identify high-risk patients receiving Intravitreal Bevacizumab injections and guide clinicians in monitoring IOP changes potentially reducing the burden of glaucoma improving the quality of life for patients receiving Intravitreal Bevacizumab injections.

## Materials and Methods

A prospective cohort study was conducted at the Ophthalmology Department of Kilimanjaro Christian Medical Centre (KCMC), a zonal referral hospital in Moshi Municipal, Kilimanjaro Region, Tanzania, from August 2023 to July 2024. The study was approved by the Kilimanjaro Christian Medical University College Research and Ethical Committee (Ref PG.83/2023), and all participants provided written informed consent before enrollment. The study was conducted in accordance with the tenets of the World Medical Association’s Declaration of Helsinki and was performed according to the Principles of Good Clinical Practice .Adult patients scheduled to receive Intravitreal Bevacizumab injections (Avastin; Genentech Inc., San Francisco, CA, USA) were recruited from the outpatient eye clinic at KCMC Hospital. Data collected included demographic information, ocular diagnosis, and indications for injection, intraocular pressure at each visit, lens status, systemic hypertension, history of YAG laser capsulotomy within six months, history of retinal surgery and number of injections.

Exclusion criteria were Glaucoma patient, Glaucoma suspect (IOP > 21 mmHg, and/or cup-to-disc ratio > 0.5), History of other Intravitreal drug injections in the same eye within 6 months (steroid, gancyclovir patient with Any pathology in either exposed or non-exposed eye that may lead to OH or secondary glaucoma and make interpretation difficult include uveitis, pseudoexfoliation syndrome, pigmentary dispersion syndrome, iridocorneal endothelial syndrome).

A minimum Sample Size of 120 was calculated using cohort sample size calculation formula using the incidence from previous study done in new York(12).

A consecutive sampling technique was used where by all patients were considered for the study until sample size was reached.

Both eye from each patient was included in the study. The injected eye was designated as the exposed eye, while the non-injected fellow eye served as the comparison, non-exposed eye. Baseline intraocular pressure (IOP) was measured before administering the Intravitreal Bevacizumab (BVM) injection using an Icare tonometer (IC100). All measurements and injection were conducted by the principal researcher, a senior ophthalmology resident in his fourth year.

Intravitreal injections were performed in a minor operating theater under aseptic conditions. The procedure involved the application of 0.5% amethocaine as topical anesthesia, followed by irrigation of the conjunctiva fornices with 5% Povidone-iodine. The lid margin and periorbital skin were disinfected using 10% Povidone-iodine. After a gentle 30-second massage, sterile skin drapes and a lid speculum were applied to prepare the site for injection. Then Intravitreal BVM injection was prepared by drawing up approximately 0.1 ml of 2.5mg BVM into 1ml insulin syringe then was injected through the infer temporal or superior temporal pars plana region at 3.5 mm from the limbus in pseudophakic and 4 mm from the limbus, in a phakic patient, Firm pressure was applied to the injection site with a cotton-tip applicator for at least 20second immediately on retrieval of the injection needle to minimize vitreous reflux.. The patient was discharged with topical dexamethasone-chloramphenicol twice a day for seven days. The IOP was measured before and immediately after injection at 5 minutes, 15 minutes, 30 minutes, and 60minutes then measured at follow-up visits of 6-week intervals for 6 months.

At 6-weekfollow-up visit, the IOP measurement was obtained before the second Injection to avoid the confounding effect from the short-term IOP rising. The mean IOP at each visit was obtained from the average of three measurements. The last IOP was measured after 6week of last injection to measure the effect of that injection.

The primary outcome measure was the proportion of eyes developing persistence IOP elevation defined as either an IOP > 21 mmHg or an increase of more than 5mmhg from the baseline.

The criteria is chosen based on a previously published definition(1,13)

Data was analyzed using STATA (Stata Corp LLC, College Station, Texas, USA) version 17). Chi-square (χ^2^) was used to determine the difference in proportion of IOP by participants’ characteristics while paired sample Student’s *t* tests was used to determine any significant difference in pre-injection and post-injection IOP at p-value of <0.05 of the study participants. The Nelson Allan estimator curves were used to show cumulative risk of ocular hypertension of the study participants during the study period and the comparison of the ocular hypertension cumulative risk between the numbers of injections. The log-rank test was used to confirm the significance difference of the ocular hypertension in the number of injections. Poisson distribution regression model was used to assess the factors associated with the ocular hypertension. Multivariate regression analysis of statistically significant potential predictors was performed by obtain the adjusted hazard ratio (AHR) to identify independent predictors of IOP changes. Variables with a P-value of < 0.05 were considered statistically associated with the event.

## Results

The median age of study participants was 62 years. The majority of participants were male (59%), and 58% had received more than three injections. A total of 16.7% of eyes had undergone YAG laser capsulotomy (**Table 1**). Diabetic macular edema (52.5%) was the leading indication for Bevacizumab injections, followed by vitreous hemorrhage (22.5%) (**Figure1**).

**Figure 1:**
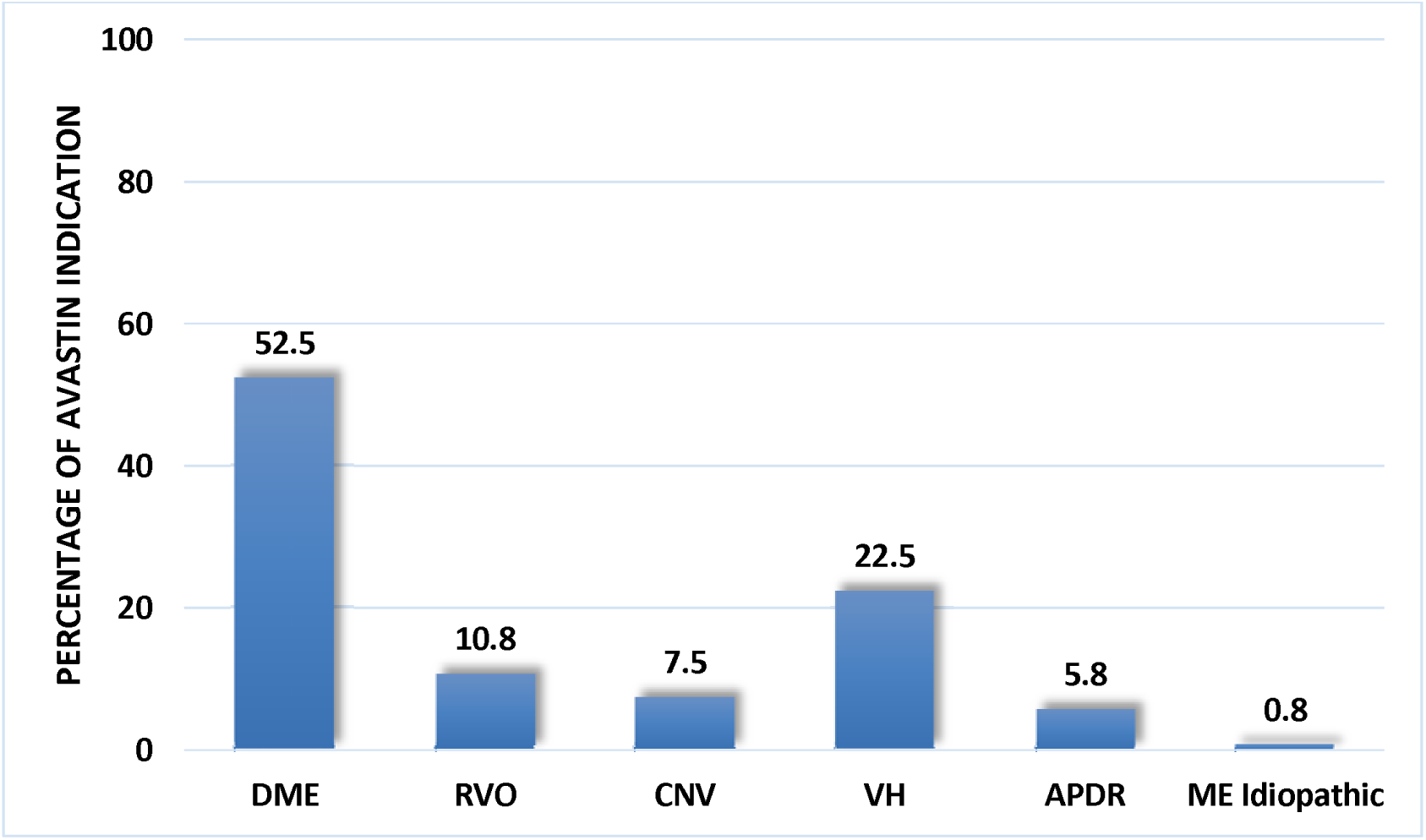
Bevacizumab indication among participants (N=120)

**Table 1 :** Participants’ background characteristics (N=120)

| Variable | Frequency | Percentage |
| --- | --- | --- |
| Sex of the participant |  |  |
| Male | 71 | 59.2 |
| Female | 49 | 40.8 |
| Age of the participant |  |  |
| ≤70 | 96 | 82.5 |
| >70 | 21 | 17.5 |
| Median(IQR) | 62(54.5-69) |  |
| Lens status |  |  |
| phakic | 95 | 79.2 |
| pseudophakic | 25 | 20.8 |
| Number of injections |  |  |
| Less than three injections | 50 | 41.7 |
| Three injections or more | 70 | 58.3 |
| YAG laser capsulotomy |  |  |
| Yes | 20 | 16.7 |
| No | 100 | 83.3 |
| **History of retina surgery** |  |  |
| No | 106 | 88.3 |
| Yes | 14 | 11.7 |
| **History of hypertension** |  |  |
| Yes | 79 | 65.8 |
| No | 41 | 34.2 |
| **RVO** |  |  |
| No | 107 | 89.2 |
| Yes | 13 | 10.8 |
| **CNV** |  |  |
| No | 111 | 92.5 |
| Yes | 9 | 7.5 |
| **VH** |  |  |
| No | 93 | 77.5 |
| Yes | 27 | 22.5 |
| **DME** |  |  |
| Yes | 57 | 47 |
| No | 63 | 52 |

The overall incidence of ocular hypertension was 15% (18/120). Where 9.2% (11/120) of study eyes were found to have persistence ocular hypertension using criteria of any increase in intraocular pressure (IOP > 21 mmHg), and 5.8 % (7/120) were identified with persistent ocular hypertension using criteria of an increase in IOP of more than 5 mmHg from baseline. Among the eyes, 1.7% (2 out of 120) showed persistent IOP elevations exceeding 30 mmHg.

Short-term intraocular pressure (IOP) spikes were observed in injected eyes following Bevacizumab administration, with distinct patterns between the first and third injections. In the first injection, 95% of eyes experienced IOP spikes, with 5% exceeding 50 mmHg at five minutes. Baseline IOP was 14 ± 2.86 mmHg, rising to 34.6 ± 5.74 mmHg five minutes post-injection, and normalizing to 14.6 ± 2.52 mmHg within 60 minutes. Most eyes (98%) returned to IOP below 21 mmHg within 30 minutes, and the rest within 60 minutes. In the third injection, 97% of eyes had IOP spikes, with 21% exceeding 50 mmHg. Baseline IOP was slightly higher at 15.8 ± 4.15 mmHg, peaking at 50 ± 16.86 mmHg and stabilizing at 17.1 ± 4.04 mmHg within 60 minutes. Normalization was slower, with 72% returning to below 21 mmHg within 30 minutes, and 28% requiring up to 60 minutes. Non-injected eyes showed minimal effects, with transient IOP increases in 3% during the first injection and 4% during the third (**Figure 2 and Figure 3**).

**Figure 2:**
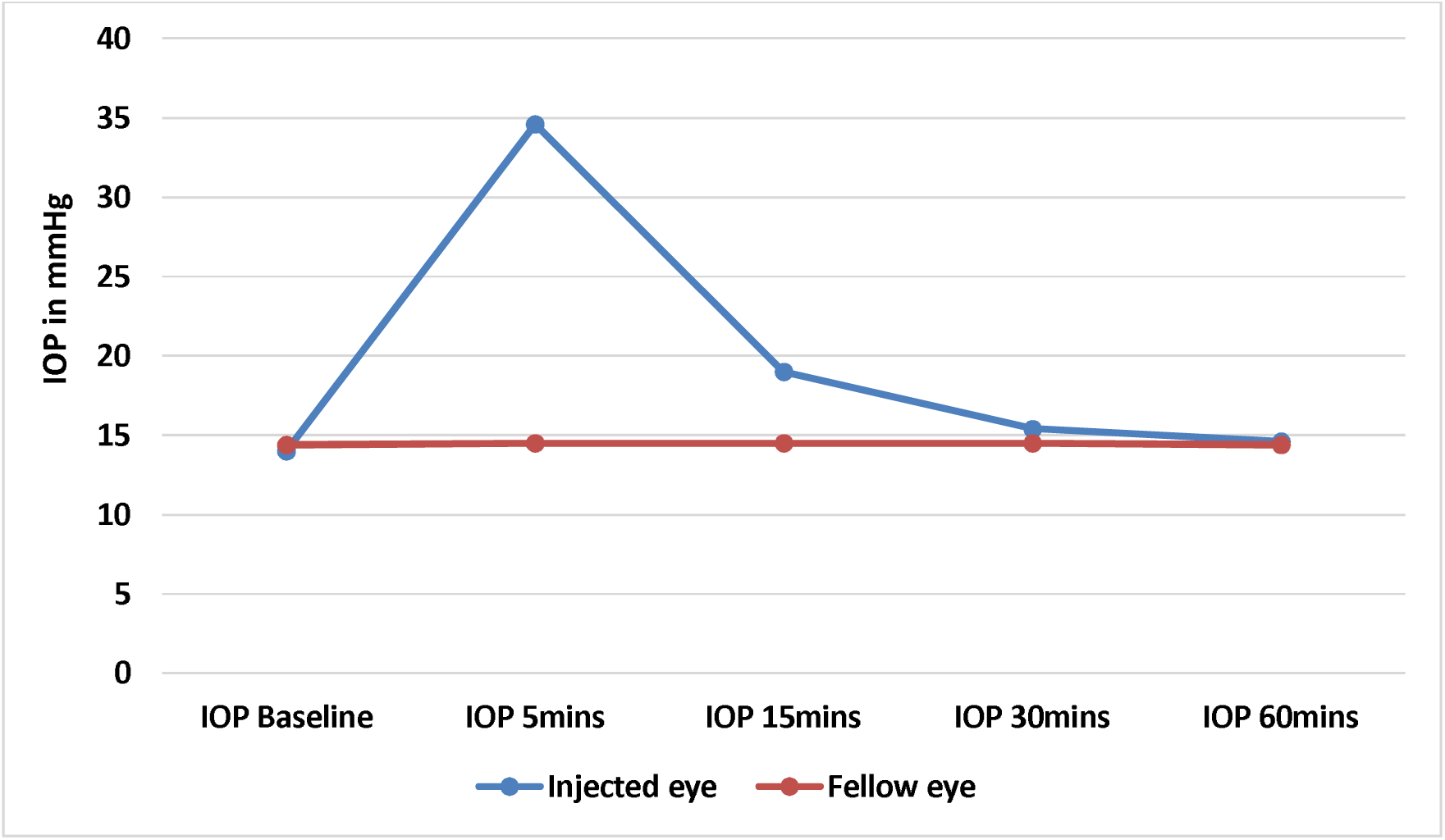
Short term pattern IOP spike after Bevacizumab injection among the study participant for first injection (Visit) (N= 120)

**Figure 3:**
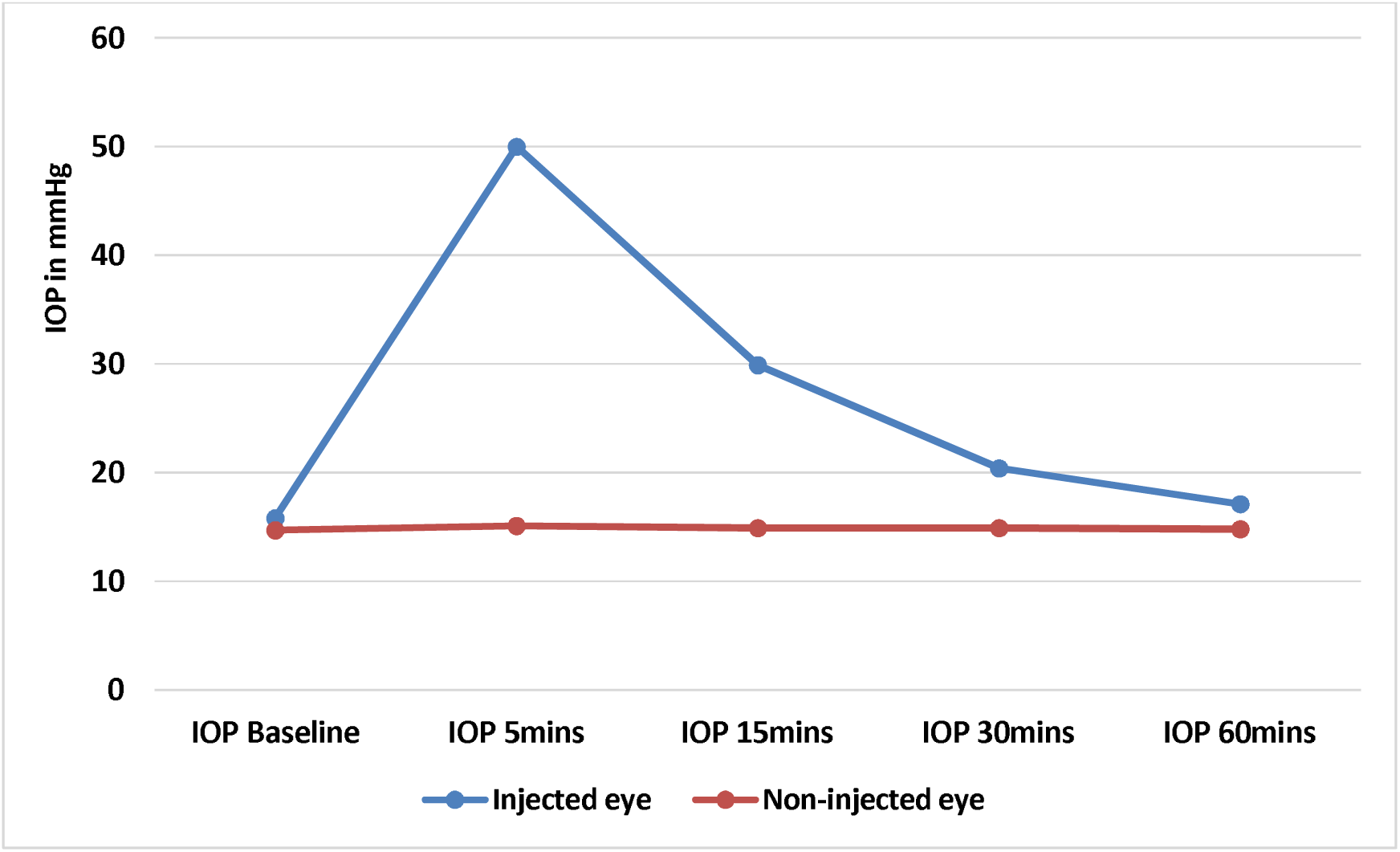
Short term pattern IOP spike after Bevacizumab injection among the study participants at third injection (12wk) (N= 120)

The mean intraocular pressure (IOP) in the injected eye progressively increased with the number of Bevacizumab injections, while the IOP in the non-injected eye remained stable throughout the study period. Before the first injection, the mean IOP was 14 ± 2.86 mmHg. After each subsequent injection, the mean IOPs at the second (6 weeks), third (12 weeks), fourth (18 weeks), and final follow-up visits were 14.7 ± 3.06 mmHg, 15.8 ± 3.56 mmHg, 16.2 ± 3.89 mmHg, and 16.4 ± 4.21 mmHg, respectively. These increases in IOP were statistically significant at each follow-up visit in the injected eye, indicating a progressive rise in pressure over time. In contrast, the non-injected eye showed no significant change in IOP during the study period (**Table 5**).

**Table 2 :**
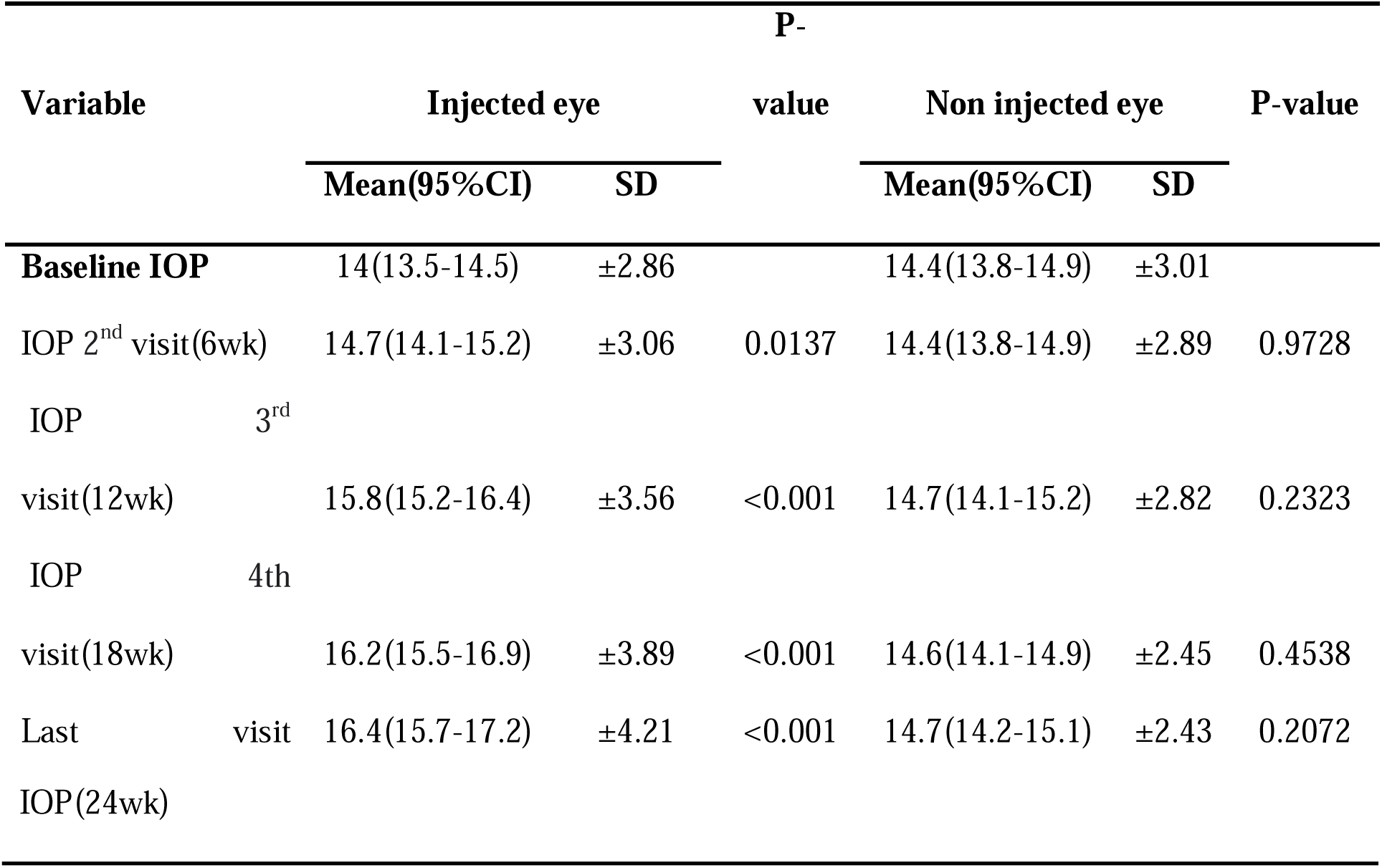
Mean Intraocular pressure at baseline and second, third, fourth, and last visit after Intravitreal injection of Bevacizumab in injected and control eye.

Ocular hypertension significantly differed by number of injection (χ^2^=11.3613, P-value=0.001) and YAG laser capsulotomy (χ^2^=7.5294, P-value=0.006) (**Table 6**).

In adjusted analysis, number of injection and history of YAG laser capsulotomy were factors that significantly associated with ocular hypertension. For every increase in number of injections, the risk of having ocular hypertension increase is 2.2 times significant higher (CHR=2.17; 95%CI (1.56-3.16) P-value< 0.001) after adjusting for YAG, while compared to participants who had YAG those without YAG, had 67% significant lower risk of having ocular hypertension (AHR=0.33; 95%CI (0.12-0.88) P-value= 0.028) after adjusting for number of number of injections age and sex (**Table 7**).

**Table 3 :** Distribution of ocular hypertension by participant’s characteristics.

| Variable | Ocular Hypertension | | $\chi^2$ | P-value |
| --- | --- | --- | --- | --- |
|  | No | Yes |  |  |
|  | n (%) | n (%) |  |  |
| **Sex** |  |  | 1.8998 | 0.168 |
| Male | 63(88.7) | 8(11.3) |  |  |
| Female | 39(79.6) | 10(20.4) |  |  |
| **Age of the participant** |  |  | 0.3271 | 0.567 |
| ≤70 | 85(85.9) | 14(14.1) |  |  |
| >70 | 17(81.0) | 4(19.0) |  |  |
| **Lens status** |  |  | 0.6192 | 0.431 |
| Phakic | 82(86.3) | 13(13.7) |  |  |
| pseudophakic | 20(80) | 5(20) |  |  |
| **Number of injections** |  |  | 11.3613 | 0.001 |
| Less than three injections | 49(98) | 1(2) |  |  |
| Three injection or more | 53(75.7) | 17(24.3) |  |  |
| **YAG laser capsulotomy.** |  |  | 7.5294 | 0.006 |
| Yes | 13(65.0) | 7(35.0) |  |  |
| No | 89(89.0) | 11(11.0) |  |  |
| **History of retina surgery** |  |  | 0.7674 | 0.381 |
| No | 89(84) | 17(16) |  |  |
| Yes | 13(92.9) | 1(7.1) |  |  |
| **History of hypertension** |  |  | 0.0065 | 0.936 |
| Yes | 67(84.8) | 12(15.2) |  |  |
| No | 35(85.4) | 6(14.6) |  |  |
| **RVO** |  |  | 0.746 | 0.388 |
| No | 92(86) | 15(14) |  |  |
| Yes | 10(76.9) | 3(23.1) |  |  |
| **CNV** |  |  | 0.398 | 0.528 |
| No | 95(85.6) | 16(14.4) |  |  |
| Yes | 7(77.8) | 2(22.2) |  |  |
| **VH** |  |  | 0.4132 | 0.52 |
| No | 78(83.9) | 15(16.1) |  |  |
| Yes | 24(88.9) | 3(11.1) |  |  |
| **DME** |  |  | 0.0793 | 0.778 |
| No | 49(85.96) | 8(14.04) |  |  |
| Yes | 53(84.13) | 10(15.87) |  |  |

**Table 4 :**
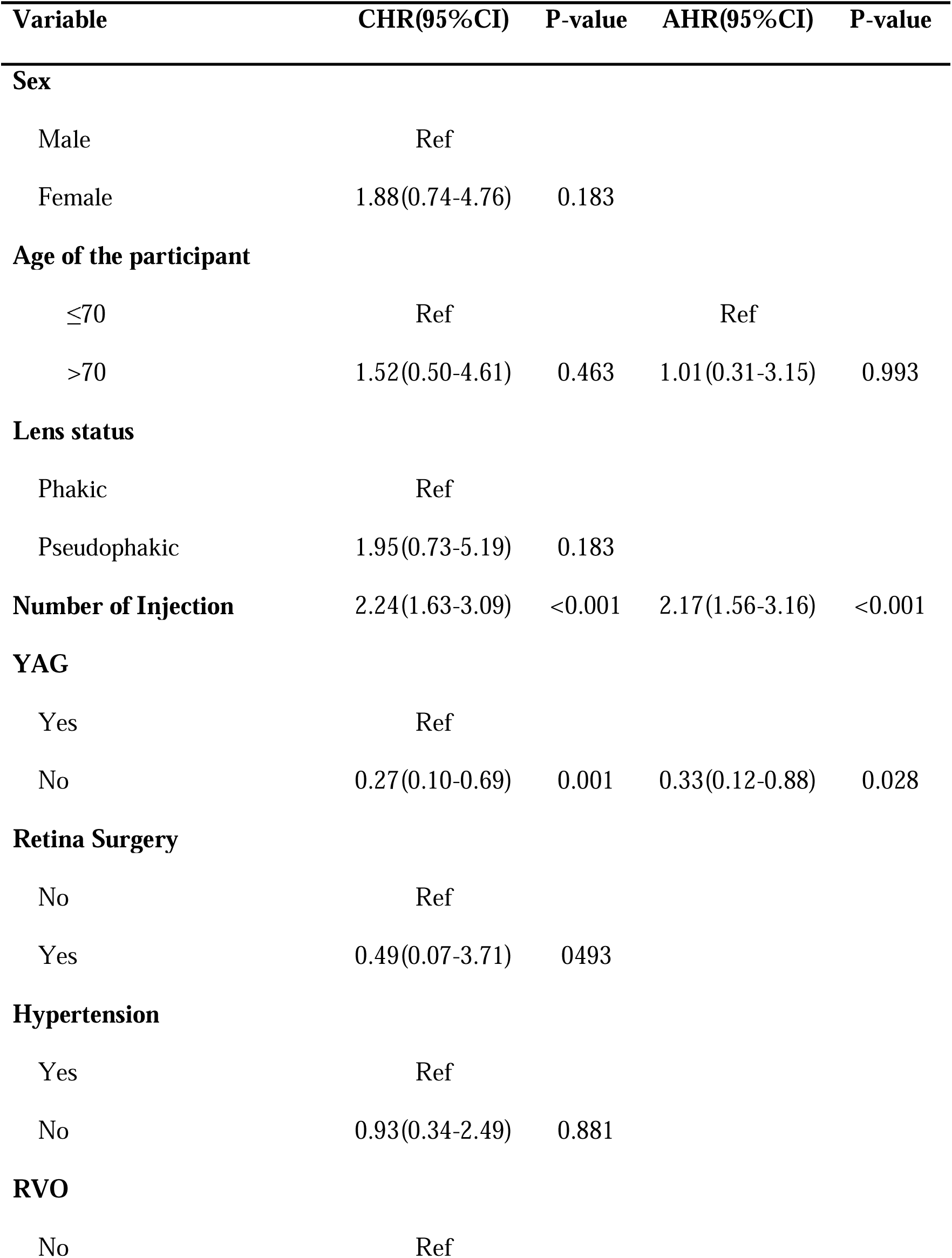
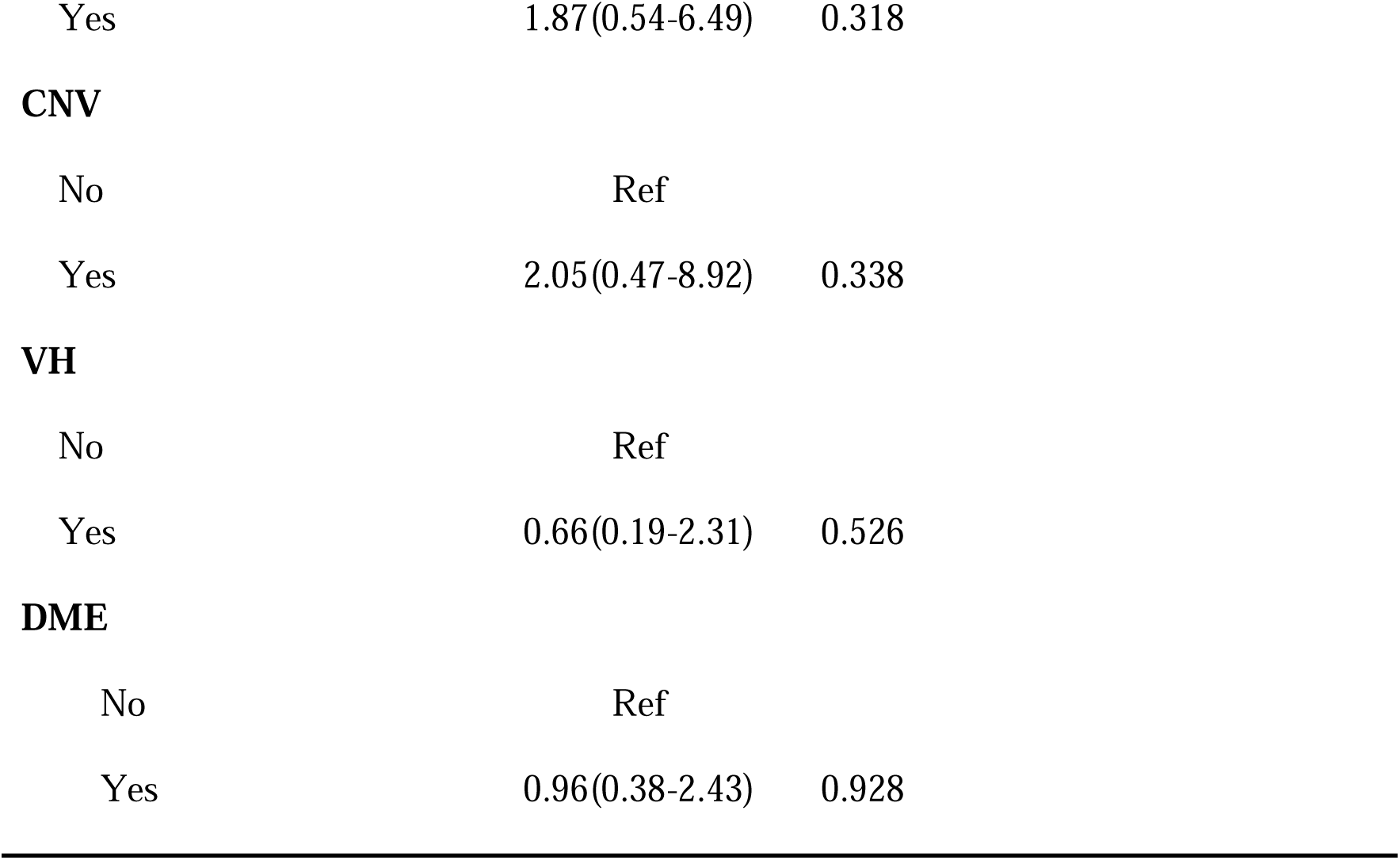
Factors associated with ocular hypertension.

| **Variable** | **CHR(95%CI)** | **P-value** | **AHR(95%CI)** | **P-value** |
| --- | --- | --- | --- | --- |
| **Sex** |  |  |  |  |
| Male | Ref |  |  |  |
| Female | 1.88(0.74-4.76) | 0.183 |  |  |
| **Age of the participant** |  |  |  |  |
| ≤70 | Ref |  | Ref |  |
| >70 | 1.52(0.50-4.61) | 0.463 | 1.01(0.31-3.15) | 0.993 |
| **Lens status** |  |  |  |  |
| Phakic | Ref |  |  |  |
| Pseudophakic | 1.95(0.73-5.19) | 0.183 |  |  |
| **Number of Injection** | 2.24(1.63-3.09) | <0.001 | 2.17(1.56-3.16) | <0.001 |
| **YAG** |  |  |  |  |
| Yes | Ref |  |  |  |
| No | 0.27(0.10-0.69) | 0.001 | 0.33(0.12-0.88) | 0.028 |
| **Retina Surgery** |  |  |  |  |
| No | Ref |  |  |  |
| Yes | 0.49(0.07-3.71) | 0.493 |  |  |
| **Hypertension** |  |  |  |  |
| Yes | Ref |  |  |  |
| No | 0.93(0.34-2.49) | 0.881 |  |  |
| **RVO** |  |  |  |  |
| No | Ref |  |  |  |

|  |  |  |
| --- | --- | --- |
| Yes | 1.87(0.54-6.49) | 0.318 |
| **CNV** |  |  |
| No | Ref |  |
| Yes | 2.05(0.47-8.92) | 0.338 |
| **VH** |  |  |
| No | Ref |  |
| Yes | 0.66(0.19-2.31) | 0.526 |
| **DME** |  |  |
| No | Ref |  |
| Yes | 0.96(0.38-2.43) | 0.928 |

## Discussion

The incidence of ocular hypertension following Bevacizumab Intravitreal injections among patients treated at KCMC Hospital from 2023 to 2024 was found to be 15%, which is notably higher than the reported average in the literature (4% to 15%;(4). Several factors contribute to the higher incidence of ocular hypertension observed at KCMC Hospital following anti-VEGF treatment. Patients likely had more advanced retinal diseases, such as diabetic retinopathy or age-related macular degeneration, which are linked to increased ocular hypertension risk. Injection protocols differed, with KCMC using larger volumes (0.1 ml of 2.5 mg Bevacizumab) and more frequent intervals (every 6 weeks) compared to the smaller doses and treat-and-extend protocols in other settings. Variations in injection techniques and post-injection care, including the use of prophylactic drugs, may also play a role. Additionally, this study employed a stricter definition for ocular hypertension, requiring both an IOP > 21 mmHg and an increase of more than 5 mmHg from baseline, likely capturing more cases than studies using a single criterion.

Comparative studies provide additional context: Belgic et al comparing Aflibercept and Ranibizumab, reported a much lower incidence of ocular 8.91%, the different can be explained by their different operational definition used (IOP≥6mmHg and/or >24mmHg on 2 or more consecutive visits) (14). Aref et al reported an 8.0% incidence of IOP increases over 60 months, lower than our study’s potentially due to different operational definition (IOP>10mmHg)(15). Maruyama-Inoue et al reported a 7% incidence of persistent IOP elevation in a Japanese cohort, lower than at KCMC, possibly due different operational definition used (IOP ≥ 25mmHg)(5).

Our findings indicate that Bevacizumab injections cause temporary increases in intraocular pressure, which become more frequent and severe with repeated treatments. While IOP levels typically return to normal within an hour, the normalization process slower with number of injection. In contrast, non-injected eyes rarely experience such changes.

The temporary IOP spike five minutes after injection is due to the mechanical response of increased intraocular volume, which may deform the lamina cribrosa and reduce ocular perfusion pressure, disturbing axonal transport. Rapid IOP normalization within 30 minutes suggests effective compensation by outflow mechanisms like the trabecular meshwork and uveoscleral pathways. The increased IOP spikes and delayed normalization after the third injection may result from cumulative effects of repeated injections, including subtle changes in ocular tissue compliance or mild inflammation.

Similarly, Dettoraki et al found that prophylactic treatment with brinzolamide-brimonidine significantly reduced IOP spikes post-injection, with the control group showing higher IOP levels at one, ten, and thirty minutes compared to the case group(10).

Similarly Lemos-Reis et al found a significant post-injection IOP rise immediately on injected eye only(16)

Conversely a study by Kato et al in retinopathy of prematurity reported IOP spike immediately after injection and at five minutes but with normalization within fifteen minutes of injection(11). Our study showed a similar rapid normalization within thirty minutes for most patients, reinforcing the transient nature of post-injection IOP spikes. The slight difference in normalization time might be due to variations study population (premature infant) and injection volume (0.625 mg/0.025 cc).

Despite these cumulative effects, the overall transient nature of the IOP spikes and the effective natural compensation mechanisms underline the safety and reliability of Intravitreal Bevacizumab injections in managing retinal conditions

With every unity increase in the number of injections was significant associated with ocular hypertension. The results of our study align with several previous studies(5,6,12,17). The consistency across these studies may be attributed to the common biological mechanism at play. This correlation can be explained by several underlying biological mechanisms, chronic inflammation and fibrosis in the trabecular meshwork (TM) and Schlemm’s canal impair aqueous humor outflow, mechanical obstruction caused by high-molecular-weight protein aggregates and Repeated injections cause micro trauma, leading to the deposition of cellular debris and protein aggregates that obstruct outflow channels.

The presence of small particles, such as immune complexes, accumulating and occluding the trabecular meshwork, has been suggested as a potential mechanism by Hoang et al(12). Additionally Liu et al reported that compounded Bevacizumab held in plastic syringes contained protein aggregates and silicone oil microdroplets, which could potentially block aqueous outflow or induce inflammatory trabeculitis(18).may also result from VEGF inhibition in the trabecular meshwork, reducing aqueous outflow facility, as proposed by Fujimoto et al(19).

Furthermore, in vitro studies by Kahook and Ammar show high Bevacizumab concentrations may damage TM cells (20).

The influence of YAG laser capsulotomy on IOP elevation is another salient finding in our study, corroborating the results of Sternfeld et al(8).Our findings indicate YAG laser capsulotomy significantly increases the risk of ocular hypertension, especially in patients receiving anti-VEGF injections. This risk arises from multifactorial mechanisms. The procedure induces transient inflammation, releasing cytokines that disrupt the trabecular meshwork. Disruption of the posterior capsule allows lens particles and inflammatory debris to enter the anterior chamber, physically obstructing the trabecular meshwork and impairing aqueous outflow. Additionally, structural changes in the lens capsule and surrounding tissues alter aqueous humor dynamics, creating inefficient drainage pathways and leading to fluid accumulation. Chronic low-grade inflammation and fibrosis from prolonged healing further exacerbate the risk of sustained IOP elevation in susceptible individuals. Conversely, Hoang et al reported no significant association between YAG capsulotomy and ocular hypertension. This divergence might be attributable to variations in study populations, differences in follow-up durations, or due to differing intervals since the capsulotomy procedure(12)

Limitations of the study include using the Icare tonometer, which isn’t the gold standard for IOP measurement, and the possible influence of topical steroid use. To mitigate these, steroids were applied less frequently (twice daily for seven days), and the mean IOP at each visit was calculated by averaging three separate measurements. Conclusion Our study at KCMC Hospital found a higher incidence of ocular hypertension following Bevacizumab Intravitreal injections compared to other studies, significant factors associated with ocular hypertension included the frequency of injections and a history of YAG laser capsulotomy. The stringent definition of ocular hypertension applied in our study likely contributed to the higher incidence. While most patients experienced rapid IOP normalization, cumulative effects from repeated injections led to delayed normalization and increased IOP spikes.

## Data Availability

All data produced in the present work are contained in the manuscrip

## Acknowledgements

This investigation partly received financial support from the Ministry of Health. Thanks for the for the Kilimanjaro Christian Medical Centre for allowing us to do this respective research

## Author Contributions

**Conceptualization:** John .A.Ambakisye.

**Formal analysis:** John .A.Ambakisye.

**Funding acquisition:** John .A.Ambakisye.

**Investigation:** John .A.Ambakisye.

**Methodology:** John .A.Ambakisye.

**Project administration:** John .A.Ambakisye.

**Writing – original draft:** John .A.Ambakisye.

**Writing – review & editing:** John .A.Ambakisye, Furahin. G.Mndeme, William Makupa

